# Vagus nerve inflammation contributes to dysautonomia in COVID-19

**DOI:** 10.1101/2023.06.14.23291320

**Authors:** Marcel S. Woo, Mohsin Shafiq, Antonia Fitzek, Matthias Dottermusch, Hermann Altmeppen, Behnam Mohammadi, Christina Mayer, Lukas C. Bal, Lukas Raich, Jakob Matschke, Susanne Krasemann, Susanne Pfefferle, Thomas Theo Brehm, Marc Lütgehetmann, Julia Schädler, Marylin M. Addo, Julian Schulze Zur Wiesch, Benjamin Ondruschka, Manuel A. Friese, Markus Glatzel

**Author notes:** **Correspondence:** (MG) or (MAF). These authors contributed equally (Manuel A. Friese, Markus Glatzel).

## Abstract

Dysautonomia has substantially impacted acute COVID-19 severity as well as symptom burden after recovery from COVID-19 (long COVID), yet the underlying causes remain unknown. Here, we show that SARS-CoV-2 is detectable in *postmortem* vagus nerve specimen together with inflammatory cell infiltration derived primarily from monocytes. This is associated with a decreased respiratory rate in non-survivors of critical COVID-19. Our data suggest that SARS-CoV-2 induces vagus nerve inflammation followed by autonomic dysfunction.

## MAIN

COVID-19 is a systemic disease affecting multiple organ systems with a wide range of acute and chronic symptoms ^1^. It is estimated that 65 million people suffer from persisting symptoms after recovery from acute COVID-19 (long COVID) with a steadily increasing prevalence ^2^. Dysautonomia is frequently reported in long COVID which can last for years or is even expected to be lifelong ^3^. Additionally, critically ill COVID-19 patients often show rapid and unpredictable vegetative deterioration which substantially contribute to the disease’s severity and lethality ^4,5^. However, the cause and mechanism of dysautonomia in COVID-19 are largely unexplored and remain elusive.

The vagus nerve is an essential component of the autonomic nervous system (ANS) and regulates critical body functions such as heart rate, digestion, and respiratory rate. Dysautonomia is observed in long COVID and other neurological diseases ^6–8^ which might be explained by vagus nerve impairment. However, the molecular underpinnings of the clinical observations of autonomic dysfunction are largely unknown. Notably, a recent histopathological study showed that the known SARS-CoV-2 entry receptors on host cells, angiotensin-converting enzyme 2 (ACE2), neuropilin 1 (NRP1), and transmembrane protease serine subtype 2 (TMPRSS2) are expressed on the vagus nerve ^9^. Therefore, we hypothesized that SARS-CoV-2 infects the vagus nerves resulting in an inflammatory response which might subsequently lead to dysautonomia during acute COVID-19 and long COVID.

To test our hypothesis, we first analyzed the vagal nuclei in the brain stem where we found an increased number of HLA-DR^+^ monocytes and CD8^+^ T cells (**Fig. 1a**). Notably, this is in line with a recent study that demonstrated a strong inflammatory response in brain stems of deceased COVID-19 patients ^10^. Next, we investigated whether the vagus nerve is directly affected by COVID-19. Therefore, we collected vagus nerves of 27 deceased patients with COVID-19 and 5 deceased controls without COVID-19 (**Fig. 1b**, demographics are shown in **extended data table 1**) and performed mRNA sequencing (**Fig. 1c, extended data fig. 1a-b**). We identified 2031 differentially up- and 42 down-regulated genes with an absolute log2 foldchange above 1. Gene ontology (GO) term analysis revealed a strong enrichment of genes regulating antiviral responses and interferon signaling (**Fig. 1d-e; extended data fig. 1c-d**), supporting the notion that vagus nerve inflammation is commonly observed in COVID-19. Next, we aimed to determine whether SARS-CoV-2 is directly detectable in vagus nerves. We performed quantitative PCR in 23 samples of deceased COVID-19 patients and detected SARS-CoV-2 RNA in all specimens which was not the case for control samples (**Fig. 1f**). To investigate whether SARS-CoV-2 RNA load correlates with vagus nerve dysfunction, we grouped the 23 vagus nerves with available SARS-CoV-2 RNA quantification into low (*n* = 6), intermediate (*n* = 9) and high (*n* = 8) SARS-CoV-2 load groups. We separately tested differential expression of all groups against controls (**Fig. 1g–i**) and performed GO term enrichment analyses. By correlating the GO terms with SARS-CoV-2 RNA load we identified GO terms that were enriched independently of SARS-CoV-2 load and GO terms that correlated with SARS-CoV-2 RNA load (**Fig. 1j**). Antigen processing, antiviral response and interferon signaling were upregulated independently of SARS-CoV-2 RNA load (**Fig. 1k**) together with the coagulation cascade and leukocyte migration (**extended data fig. 1e**). Notably, complement activation and regulation of apoptotic cell clearance showed a direct correlation with viral RNA load (**Fig. 1l**). In contrast, we found that genes regulating trans-synaptic signaling or assembly of the neuronal transport machinery had an inverse correlation with SARS-CoV-2 RNA load (**Fig. 1m**) suggesting dose-dependent axonal dysfunction.

**Fig. 1.**
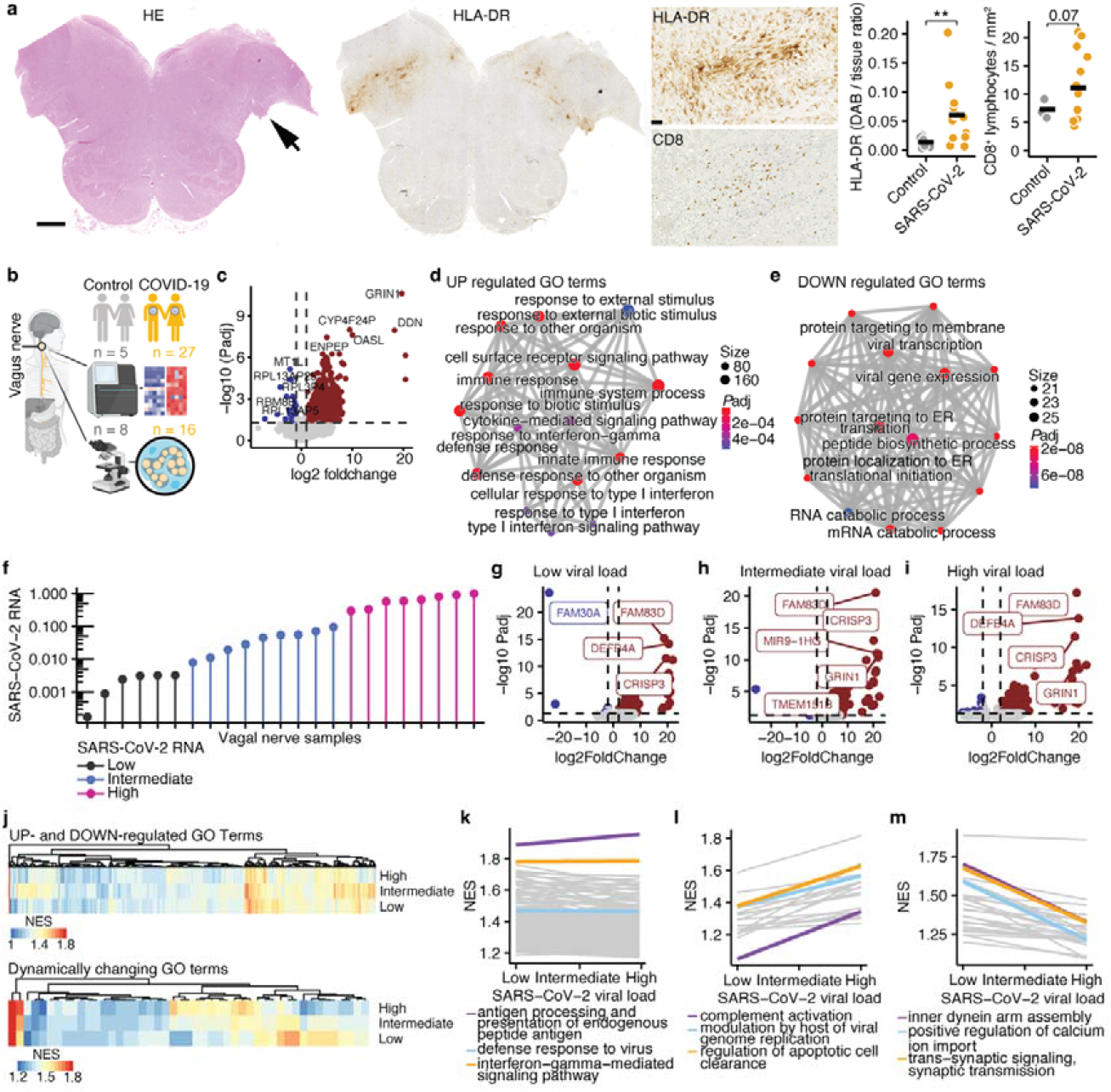
Inflammation of the vagus nerve is commonly observed in COVID-19. **(a)** H&E staining (left) and HLA-DR staining (middle) with arrow indicating the entry of the vagal nerve. Scale bar shows 2 mm. Higher magnification of vagal nerve entry into the brain stem of HLA-DR (middle, top) and CD8 (middle, bottom) staining and quantification. Scale bar shows 100 µm. **(b)** Graphical summary of experimental setup. **(c)** Volcano plot depicting all identified genes. Genes with FDR-adjusted *P* value < 0.05 and log2 foldchange > 1 are labeled in red and < –1 in blue. Top 5 most upregulated and downregulated genes are labeled. **(d)** GO term analysis of top 200 differentially upregulated genes. Color shows *P*adj and size number of genes included in the respective GO term. **(e)** GO term analysis of top 200 differentially downregulated genes. Color shows FDR-adjusted *P* value and size number of genes included in the respective GO term. **(f)** SARS-CoV-2 RNA levels determined by qPCR in 23 available vagus nerve samples grouped into low, intermediate, and high groups. Logarithmic scale is shown on y-axis. **(g–i)** Volcano plots of differential expression analyses between low (g), intermediate (h), and high (i) viral load groups against controls. **(j)** Heatmaps of GO terms that are up- or downregulated independently of SARS-CoV-2 viral load (top) or correlate with SARS-CoV-2 viral load (down) determined by generalized linear model corrected for age and sex. **(k)** GO terms that are significantly enriched independently of SARS-CoV-2 viral load. **(l)** GO terms that are positively associated with SARS-CoV-2 viral load. **(m)** GO terms that are negatively associated with SARS-CoV-2 viral load.

Next, we aimed to identify the SARS-CoV-2-dependent cell type composition and activation in vagus nerves. We performed weighted gene correlation network analysis (WGCNA; **Fig. 2a, extended data fig. 2a**) and identified modules which could be assigned to unique cell types by marker gene expression. By using generalized linear models, we identified modules which were enriched in COVID-19. These included NK cell modules, a monocyte module as well as several endothelial cell and neuronal modules (**Fig. 2b**), suggesting cell type-specific inflammatory responses in vagus nerves of COVID-19 patients. Since we identified interferon signaling as overarching theme independently of SARS-CoV-2 viral load we first interrogated this theme specifically and respective GO terms in different cell types. In accordance, almost all cell types showed a strong positive enrichment for interferon signaling, including neurons and endothelial cells, further supporting that the interferon cascade critically contributes to vagus nerve dysfunction in COVID-19. Notably, we identified one B cell subtype which was strongly de-enriched for interferon pathways, which was not differently regulated in COVID-19 (**Fig. 2c**). Next, we focused on modules which were stronger enriched in COVID-19 in comparison to controls (**extended data fig. 2b**). We found that monocytes (“orange”) showed an activated transcriptional phenotype (**Fig. 2d**) whereas we observed stress responses in endothelial cells (“turquoise”; **Fig. 2e**) and Schwann cells (“brown”; **Fig. 2f**) including regulation of autophagy, endoplasmic reticulum (ER) stress and proteasomal catabolism. Similarly, in the neuronal modules that were significantly enriched in COVID-19 we observed stress responses such as the unfolded protein response and ER stress response (“saddle brown”; **Fig. 2g**).

**Fig. 2.**
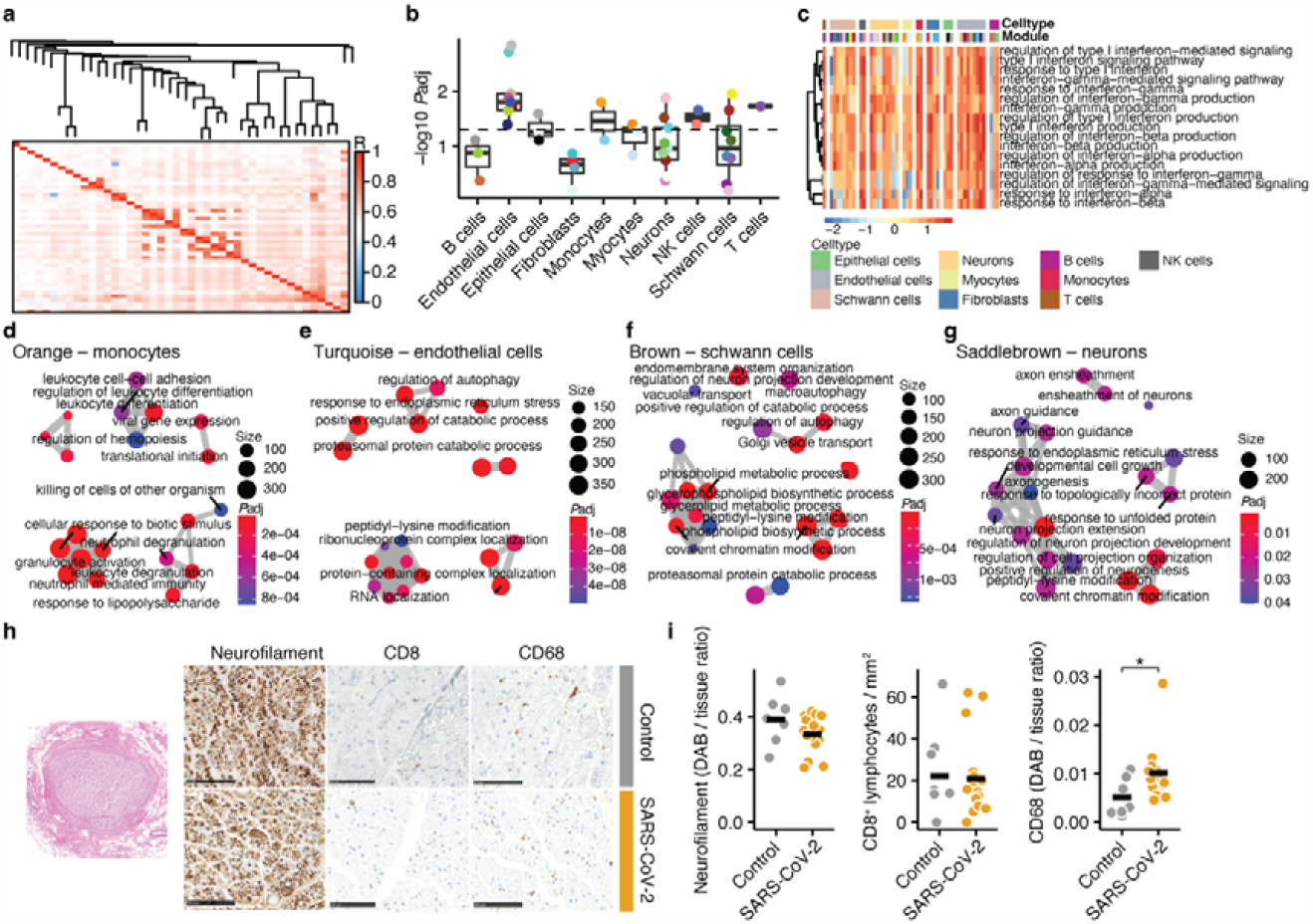
Cell type-dependent inflammatory responses in vagus nerves of COVID-19 patients. **(a)** Network heatmap plot of weighted gene correlated network analysis (WGCNA) for module identification in all samples. (b) Modules separated by clearly identified cell types through enrichment analysis with PanglaoDB. Modularity between control and COVID-19 samples were compared within each module. *P* < 0.05 shows significantly stronger enrichment of COVID-19 samples. Dashed line represents *P* = 0.05. Color represents module. **(c)** Heatmap depicting the enrichment of interferon signaling and response GO terms in all identified modules. Cell types and modules are separately labeled as individual colors. Color range shows row z-score. Non-significant enrichment is depicted as grey tile. **(d–g)** Emap plots of GO term enrichment analyses in the orange monocyte (d), turquoise endothelial cell (e), brown Schwann cell (f), and saddlebrown neuronal (g) modules. Interferon signaling GO terms were excluded. Color shows FDR-adjusted *P* value. Size shows number of genes included in the GO term. **(h)** Representative images of neurofilament, CD8, and CD68 in vagus nerves of control (top) and COVID-19 (bottom) samples. Scale bar shows 50 µm. **(i)** Comparison of neurofilament (DAB / tissue intensity ratio), CD8^+^ T cells per mm^2^ and CD68 (DAB / tissue intensity ratio) between control (*n* = 8, except for CD68 where 1 outlier was removed since it was > 3.5x z-score of all samples) and COVID-19 samples (*n* = 16). Student’s *t*-test was used for statistical comparison. \**P* < 0.05.

We validated the transcriptional findings by immunohistochemical analyses of vagus nerves from deceased COVID-19 patients (**Fig. 2h**) and found an increased number of CD68^+^ monocytes, whereas the extent of CD8^+^ T cells was similar in comparison to control samples. Of note, we did not find differences in axonal damage assessed by quantifying neurofilament, indicating that SARS-CoV-2-induced inflammation and neuronal stress responses do not lead to overt degeneration of the vagal neurons (**Fig. 2i**).

Lastly, we tested the hypothesis that vagus nerve dysfunction predicts disease severity and mortality. We investigated a cohort of 323 patients admitted to emergency room with acute COVID-19 between February 13, 2020 and August 15, 2022 (patient demographics are summarized in **extended data table 2**) and analyzed vital signs as well as laboratory parameters of inflammation. We separated the patients into five disease severities (mild, moderate, severe, critical, lethal) which was reflected by oxygen saturation (**Fig. 3a**) and inflammatory markers (**Fig. 3b-c, extended data fig. 2c**). The spontaneous respiratory rate at admission was used as a biomarker for vagus nerve dysfunction due to the well-documented observations that stimulating the vagus nerve leads to an increased respiratory rate ^11^. As expected, the respiratory rate steadily increased with disease severity. However, when looking at patients with critical COVID-19, we found that the respiratory rate had increased in survivors but not in non-survivors (**Fig. 3d**). Additionally, reduced respiratory rate was the strongest predictor for mortality in critical COVID-19 (**Fig. 3e**, summary statistics is provided in **extended data table 3**), supporting our hypothesis of vagus nerve dysfunction in COVID-19. Next, we asked whether the respiratory rate is regulated by physiological modulators of respiration. Therefore, we analyzed the associations between the respiratory rate and blood biomarkers of respiratory acidosis. Our analysis revealed that an increased respiratory rate was significantly associated with elevated oxygen levels in the blood (**Fig. 3f**) but decoupled from physiologic stimulators of respiration, such as carbon dioxide levels and acidosis in the blood (**Fig. 3g-h**). Confirmatory, respiratory rate did not correlate with inflammation in survivors and non-survivors of critical COVID-19 (**Fig. 3i, extended data fig. 2d**, summary statistics is provided in **extended data table 4**).

**Fig. 3.**
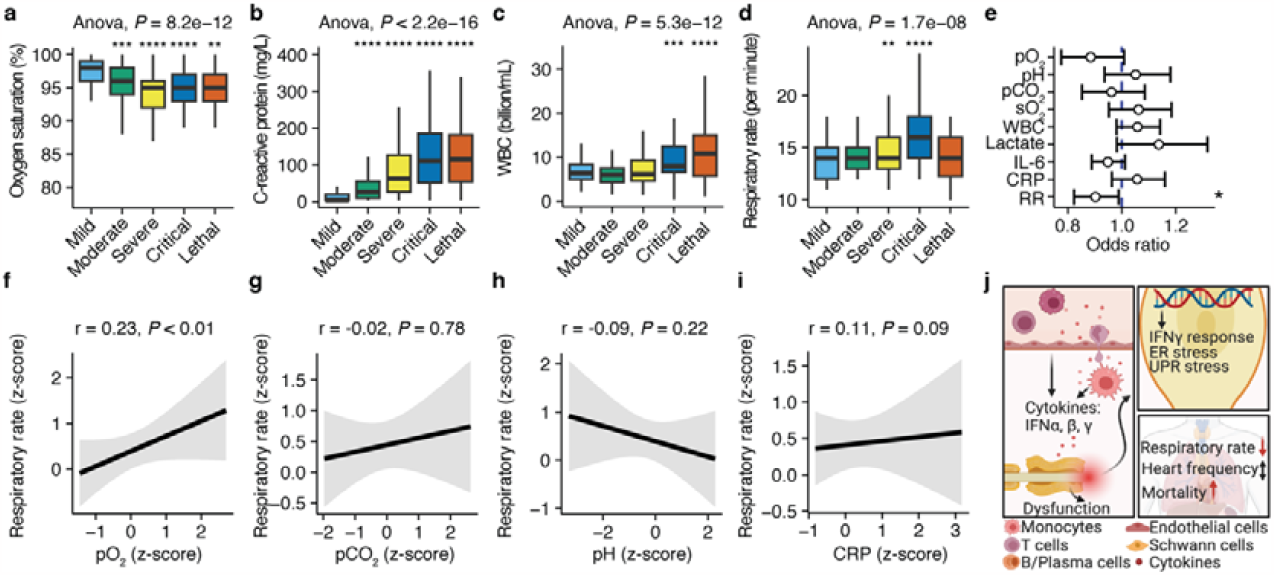
Maladaptive respiratory rate predicts lethal outcome in COVID-19. **(a–d)** Comparisons of oxygen saturation (in percent; a), C-reactive protein (mg L^−1^; b), white blood cell count (billion mL^−1^; c), and respiratory rate (per minute; d) in patients with mild, moderate, critical, and lethal COVID-19. One-way ANOVA was performed for testing group differences. *T*-test with FDR-correction for multiple comparisons against mild COVID-19 group was performed. \**P* < 0.05, \*\**P* < 0.01, \*\*\**P* < 0.001, \*\*\*\**P* < 0.0001. (e) Odds ratio analysis corrected for age and sex to predict lethal outcome in critically ill patients. pH, venous carbon dioxide levels (pCO_2_), peripheral oxygen saturation (sO_2_), white blood cell count (WBC), lactate, interleukin-6 (IL-6), C-reactive protein (CRP) and respiratory rate (RR) were tested as predictors. Odds ratio and 95% confidence intervals are shown. \**P* < 0.05. **(f–i)** Correlation analyses of z-transformed respiratory rate and z-transformed venous oxygen level (pO_2_; f), venous carbon dioxide level (pCO_2_; g), pH (h), and C-reactive protein (CRP; i). Spearman correlation analysis was performed. Correlation coefficients and *P* values are provided in the figure. **(j)** Graphical summary of vagus nerve inflammation in COVID-19.

Overall, our data show that the presence of SARS-CoV-2 RNA in the vagus nerve as well as vagus nerve inflammation and dysfunction are widespread in COVID-19 (graphical summary provided in **Fig. 3i**). This is in line with large observational studies reporting dysautonomia as a common symptom of long COVID ^12,13^. Here, by detecting SARS-CoV-2 RNA we demonstrate that SARS-CoV-2 is able to infect vagus nerves which in turn causes functional impairment and decoupling from physiological modulators of respiration that predicts lethal disease outcome. The vagus nerve could be particularly susceptible to SARS-CoV-2 infection, as it is directly innervating the respiratory tract, which is the primary site of infection. Other entry routes could be direct axonal infection, since the receptors of SARS-CoV-2 are highly abundant on the vagus nerve, or through endothelial cells similar to CNS infection ^14,15^. This is supported by our findings of strong endothelial inflammatory responses in vagus nerves of COVID-19 patients similar to the transcriptional profiles of the neurovascular units in the CNS ^16,17^. Thereby, the vagus nerve might serve as additional gateway for SARS-CoV-2 infiltration of the brain stem and CNS ^10^. Across different cell types, SARS-CoV-2 induced a strong interferon response independent of the viral load. This has been reported for several immune subsets and increased interferon levels in the serum are highly predictive for systemic inflammation ^18,19^. Notably, SARS-CoV-2 RNA load was associated with complement activation and negatively correlated with assembly of the neuronal transport machinery. The complement system is an important mediator of neuronal signaling and its crucial role in neuroinflammation in the CNS has been demonstrated, for instance in neuromyelitis optica, multiple sclerosis and Alzheimer’s disease ^20,21^. Further mechanistic studies are required to evaluate the role of the complement system in damaging the ANS and especially the vagus nerve. Due to high prevalence of dysautonomia in long COVID and numerous other neurological and infectious diseases, our discovery of vagus nerve inflammation sheds light on the pathophysiology of autonomic dysfunction.

## METHODS

### Human samples

Autopsies were performed at the Institute of Legal Medicine of the University Medical Center Hamburg-Eppendorf, Germany. Use of *postmortem* human tissue after conclusion of diagnostic procedures were reviewed and approved by the institutional review board of the independent ethics committee of the Hamburg Chamber of Physicians (protocol nos. PV7311, 2020-10353-BO-ff, and PV5034). Vagal nerve tissues were used for this study. For COVID-19, *postmortem* tissue samples were used; for non-COVID-19 controls, we used samples from individuals with other cause of death. A summary of included samples is provided in **extended data table 1**.

### Histopathology of Human tissue

Formalin-fixed paraffin embedded tissue (FFPE) samples from the vagal nerve were processed and stained with hematoxylin and eosin following standard laboratory procedures. Furthermore, immunohistochemistry with antibodies to human leukocyte antigen DR, (HLA-DP, DQ and DR) (1:200, mouse clone CR3/43; DakoCytomation), Cluster of Differentiation 68 (CD68) (1:100, clone PG-M1, DakoCytomation, Glostrup, Denmark), human Cluster of Differentiation 8 (CD8) (1:100, clone SP239, Spring Bioscience, Pleasanton, USA) was performed on a Ventana benchmark XT autostainer following the manufacturer’
ss recommendations. The quality of the immunohistochemical staining was assessed by on-slide positive controls for all antibodies.

### RNA sequencing

Total RNA was isolated from the fresh frozen tissues using RNeasy mini kit (Cat. No. 74106; Qiagen, Hilden, Germany), following the manufacturer’s suggested protocol. RNA sequencing libraries were prepared using the TruSeq stranded mRNA Library Prep Kit (Illumina) according to the manufacturer’s manual (document 1000000040498 v00) with a minimum total RNA input of 100 ng per sample. Libraries were pooled and sequenced on a NovaSeq 6000 sequencer (Illumina) generating 50 bp paired end reads. The reads were aligned to the Ensembl human reference genome (GRCh38) using STAR v.2.4 ^22^ with default parameters. The overlap with annotated gene loci was counted with featureCounts v.1.5.1 ^23^. Differential expression analysis was performed with DESeq2 (v.3.12) ^24^ calling genes with a minimal 2-fold change and false discovery rate (FDR)-adjusted *P* < 0.05 differentially expressed. Gene lists were annotated using biomaRt (v.4.0) ^25^. Gene set enrichment analysis (GSEA) was performed using the *clusterProfiler* package ^26^.

### Weighted gene correlated network analysis (WGCNA)

We performed WGCNA to identify co-correlating modules that could be assigned to cell types using the *WGCNA* package in R (v4.0.3) ^27^. Only genes that were detectable in more than 3 samples were used. Modules were constructed by signed Pearson’s correlation using a soft threshold power of 12 corresponding to a scale free model fit > 0.9 with a minimum module size of 20 genes per module. Subsequent cell type annotation was performed by enrichment analysis using markers provided by the PanglaoDB ^28^. FDR-corrected *P* values < 0.05 were considered statistically significant. Cell type annotation was further confirmed by gene set enrichment analysis (GSEA) of the modules using *clusterProfiler* ^26^. Differences in module enrichment of different genotypes was determined by comparing the module eigenvalues per timepoint using generalized linear model corrected for age and sex.

### Human clinical data

To assess a clinical phenotype of vagus nerve dysfunction we included all patients who were admitted to the University Medical Center Hamburg-Eppendorf from February 13, 2020 until August 15, 2022 with COVID-19. All participants gave written consent in this study that was approved by the local ethics board. All patients with COVID-19 where positive for SARS-CoV-2 by PCR or had SARS-CoV-2 specific antibody titers. The severity of COVID-19 into mild (no/mild symptoms without oxygen support), moderate (oxygen supplementation required at < 2 L min^−1^), severe (oxygen supplementation required at > 2 L min^−1^ OR complication), critical (submission to intensive care unit), and lethal disease courses was classified based on the most affected pathologies per day during the hospital stay. Patient demographics are summarized in **extended data table 2**. For comparisons of vital signs, we used the first recorded data on day of admission where no possible confounding treatments were started that could bias the analyses. Additionally, all patients who were admitted with invasive ventilation to our hospital were excluded since in these cases the respiratory rate was externally controlled.

### Statistical analysis

All statistical analyses and visualizations were performed within the R environment (v4.0.3). The detailed analyses of RNA-sequencing data are described above. For immunohistochemical quantification we compare the 2 groups by Student’s *t*-test. For clinical data, we first performed 1-way ANOVA to analyze variance on group levels and subsequently *t*-test with FDR-correction for multiple comparisons. Modularity between controls and SARS-CoV-2 positive samples were compared for each identified module by Mann-Whitney-U test. Odds ratios for lethal disease outcome were calculated in patients with critical COVID-19 by generalized mixed models with correction for age and sex. For correlation analyses we first performed z-scaling to account for outliers and used Spearman-correlation for testing significance. *P* values < 0.05 were considered significant.

## Data Availability

The RNA-sequencing data is available under the GEO accession number c. All other data is available from the corresponding author upon reasonable request.

## DATA AVAILABILITY

The RNA-sequencing data is available under the GEO accession number GSE233732. All other data is available from the corresponding author upon reasonable request.

## CODE AVAILABILITY

The code is available from the corresponding author upon reasonable request.

## ACKNOWLEDGEMENTS

We like to thank members of the Glatzel and Friese laboratories for helpful discussions. Graphical abstract and summary were generated with Biorender.

## AUTHOR CONTRIBUTIONS

MSW, MG, MAF conceptualized the study. MSW performed most analyses. MS, HCA prepared the RNA for mRNA sequencing. MS, HCA, MD, BM, JM, SK performed analysis of histopathology data. AF, BO, JS helped with preparations and sampling of vagus nerves. AF, MS, HCA managed/handled frozen tissue samples. CM, LCB, LR, MMA, JSZW helped with acquisition and analysis of clinical data. SP, ML performed SARS-CoV-2 PCRs. MSW, MAF and MG supervised the study and wrote the first version of the manuscript. MAF and MG funded the study. All authors approved the final version of the manuscript.

## EXTENDED DATA FIGURES

**Extended data fig. 1.**
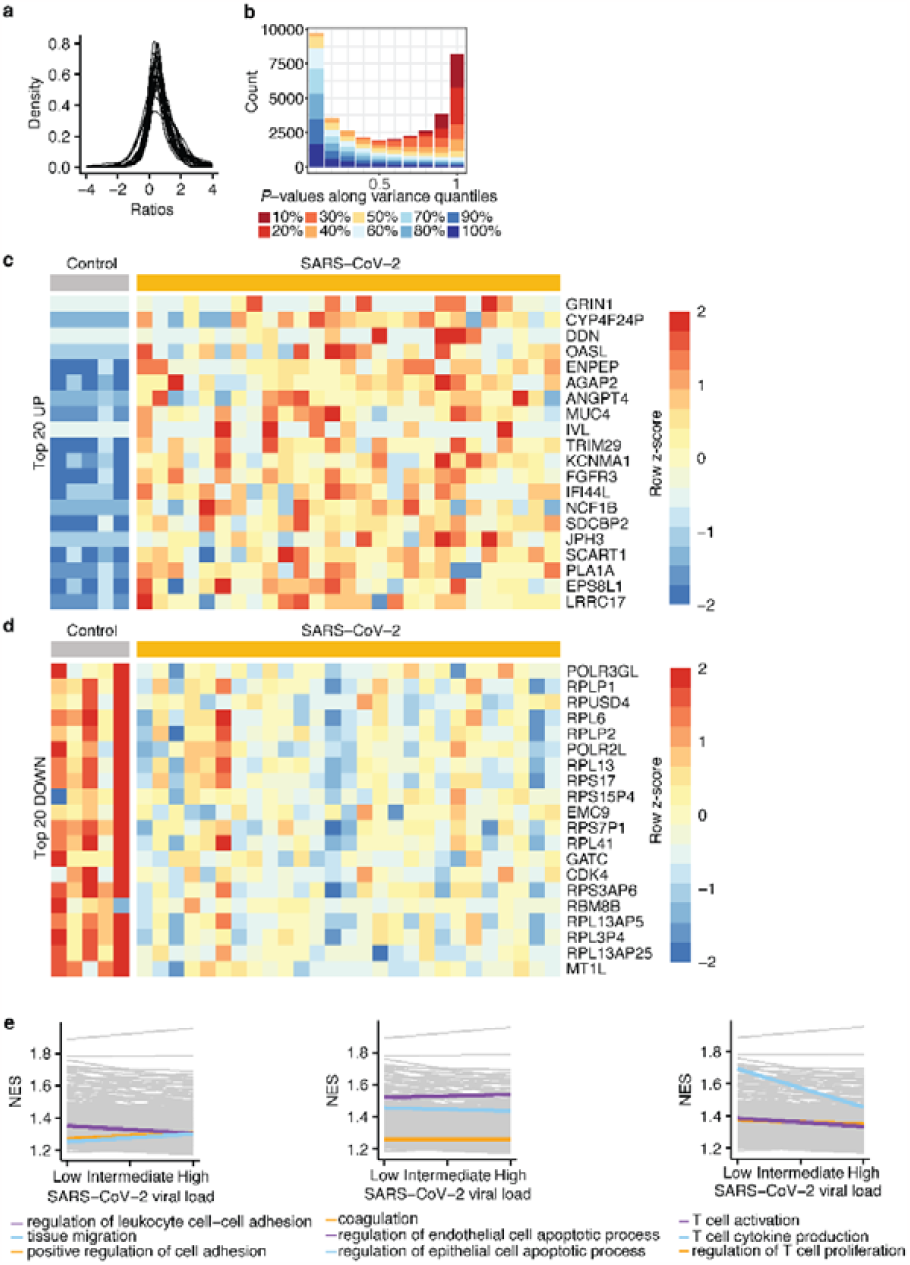
RNA sequencing of vagus nerves. **(a–b)** Quality control of RNA sequencing experiments. Density distribution of detected genes (a) and *P* values of genes along the variance quantiles (b). **(c–d)** Top 20 upregulated (c) and downregulated (d) genes in vagus nerves of COVID-19 patients in comparison to controls. Color shows row z-score. **(e)** GO terms that are significantly upregulated independently of SARS-CoV-2 viral load.

**Extended data fig. 2.**
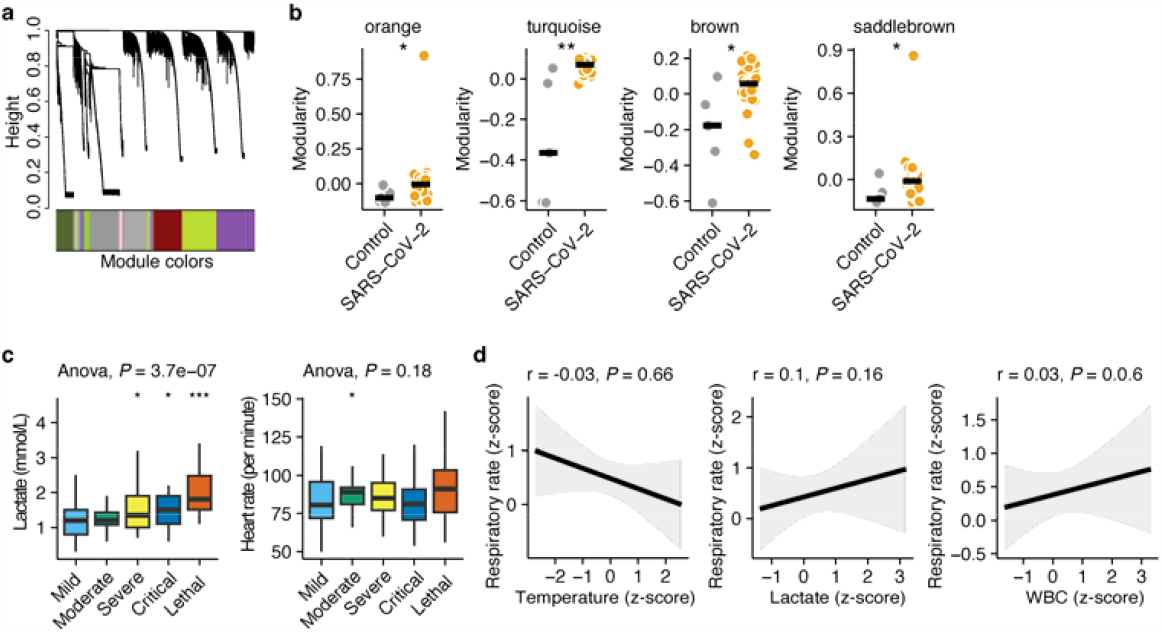
Respiratory rate is decoupled from inflammation in COVID-19. **(a)** Dendrogram of weighted gene correlated network analysis (WGCNA) for module detection. **(b)** Comparison of module enrichment by COVID-19 samples in orange, turquoise, brown and saddlebrown modules. Wilcoxon-test was used for statistical comparison. \**P* < 0.05, \*\**P* < 0.001. **(c)** Comparison of lactate and heart rate in patients with mild, moderate, critical, and lethal COVID-19. One-way ANOVA was performed for testing group differences. *T*-test with FDR-correction for multiple comparisons against mild COVID-19 group was performed. \**P* < 0.05, \*\**P* < 0.01, \*\*\**P* < 0.001. **(d)** Correlation analyses of z-transformed respiratory rate and z-transformed temperature, lactate, and white blood cell count (WBC). Spearman correlation analysis was performed. Correlation coefficients and *P*-values are provided in the figure.

## EXTENDED DATA TABLES

**Extended data table 1.**
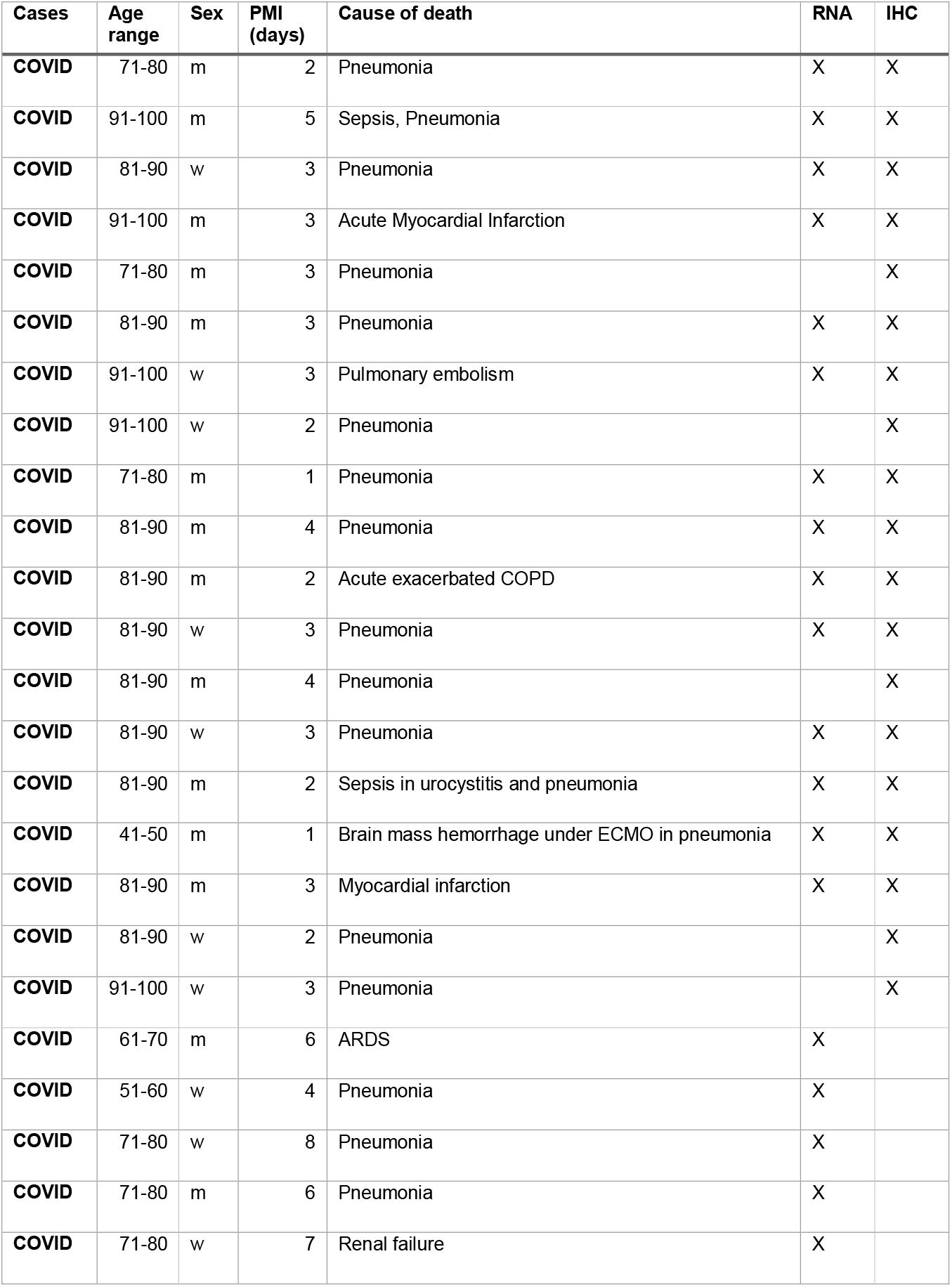
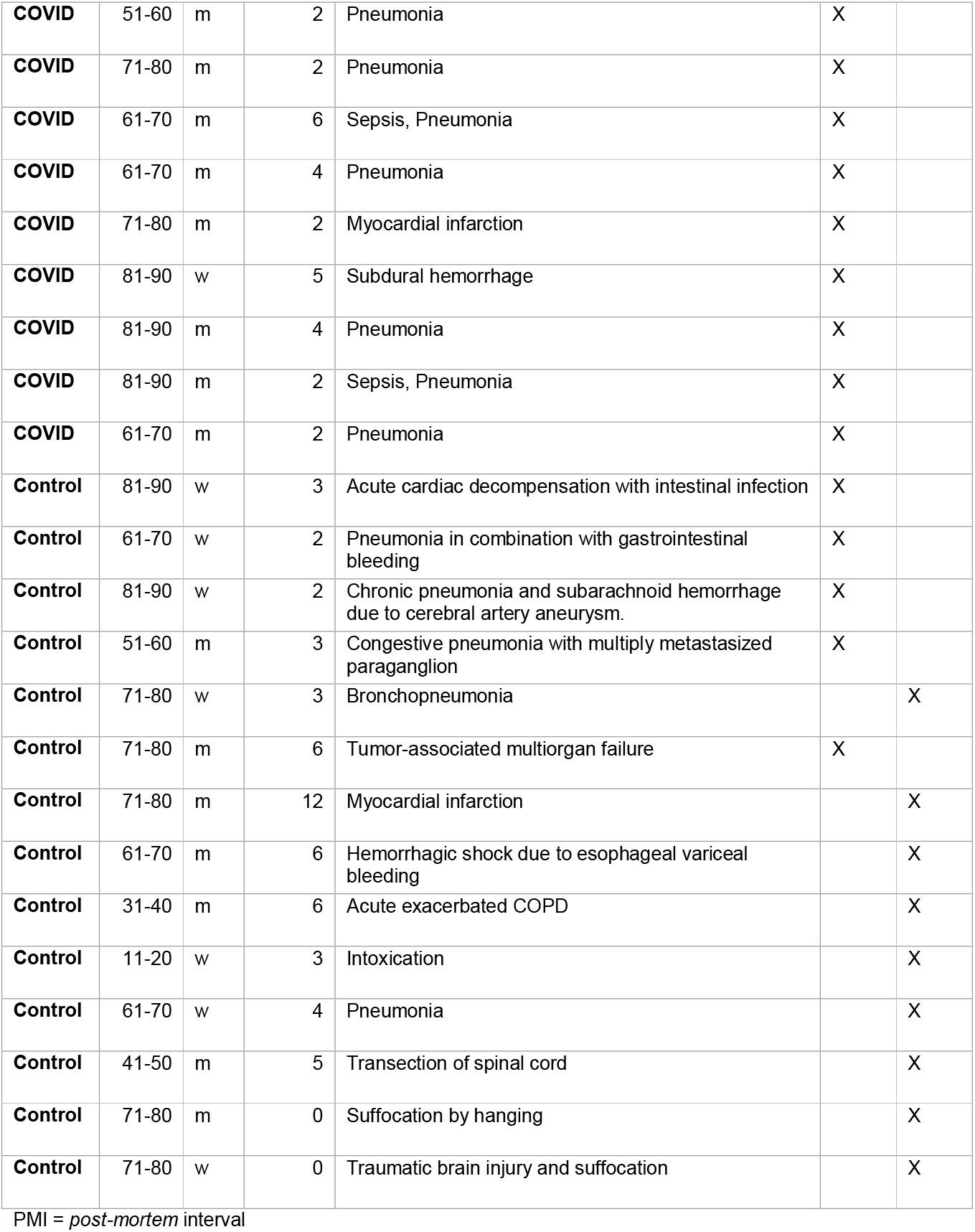
Patients’ demographics for vagus nerve analyses.

| Cases | Age range | Sex | PMI (days) | Cause of death | RNA | IHC |
| --- | --- | --- | --- | --- | --- | --- |
| COVID | 71-80 | m | 2 | Pneumonia | X | X |
| COVID | 91-100 | m | 5 | Sepsis, Pneumonia | X | X |
| COVID | 81-90 | w | 3 | Pneumonia | X | X |
| COVID | 91-100 | m | 3 | Acute Myocardial Infarction | X | X |
| COVID | 71-80 | m | 3 | Pneumonia |  | X |
| COVID | 81-90 | m | 3 | Pneumonia | X | X |
| COVID | 91-100 | w | 3 | Pulmonary embolism | X | X |
| COVID | 91-100 | w | 2 | Pneumonia |  | X |
| COVID | 71-80 | m | 1 | Pneumonia | X | X |
| COVID | 81-90 | m | 4 | Pneumonia | X | X |
| COVID | 81-90 | m | 2 | Acute exacerbated COPD | X | X |
| COVID | 81-90 | w | 3 | Pneumonia | X | X |
| COVID | 81-90 | m | 4 | Pneumonia |  | X |
| COVID | 81-90 | w | 3 | Pneumonia | X | X |
| COVID | 81-90 | m | 2 | Sepsis in urocystitis and pneumonia | X | X |
| COVID | 41-50 | m | 1 | Brain mass hemorrhage under ECMO in pneumonia | X | X |
| COVID | 81-90 | m | 3 | Myocardial infarction | X | X |
| COVID | 81-90 | w | 2 | Pneumonia |  | X |
| COVID | 91-100 | w | 3 | Pneumonia |  | X |
| COVID | 61-70 | m | 6 | ARDS | X |  |
| COVID | 51-60 | w | 4 | Pneumonia | X |  |
| COVID | 71-80 | w | 8 | Pneumonia | X |  |
| COVID | 71-80 | m | 6 | Pneumonia | X |  |
| COVID | 71-80 | w | 7 | Renal failure | X |  |
| **COVID** | 51-60 | m | 2 | Pneumonia | X |  |
| **COVID** | 71-80 | m | 2 | Pneumonia | X |  |
| **COVID** | 61-70 | m | 6 | Sepsis, Pneumonia | X |  |
| **COVID** | 61-70 | m | 4 | Pneumonia | X |  |
| **COVID** | 71-80 | m | 2 | Myocardial infarction | X |  |
| **COVID** | 81-90 | w | 5 | Subdural hemorrhage | X |  |
| **COVID** | 81-90 | m | 4 | Pneumonia | X |  |
| **COVID** | 81-90 | m | 2 | Sepsis, Pneumonia | X |  |
| **COVID** | 61-70 | m | 2 | Pneumonia | X |  |
| **Control** | 81-90 | w | 3 | Acute cardiac decompensation with intestinal infection | X |  |
| **Control** | 61-70 | w | 2 | Pneumonia in combination with gastrointestinal bleeding | X |  |
| **Control** | 81-90 | w | 2 | Chronic pneumonia and subarachnoid hemorrhage due to cerebral artery aneurysm. | X |  |
| **Control** | 51-60 | m | 3 | Congestive pneumonia with multiply metastasized paraganglion | X |  |
| **Control** | 71-80 | w | 3 | Bronchopneumonia |  | X |
| **Control** | 71-80 | m | 6 | Tumor-associated multiorgan failure | X |  |
| **Control** | 71-80 | m | 12 | Myocardial infarction |  | X |
| **Control** | 61-70 | m | 6 | Hemorrhagic shock due to esophageal variceal bleeding |  | X |
| **Control** | 31-40 | m | 6 | Acute exacerbated COPD |  | X |
| **Control** | 11-20 | w | 3 | Intoxication |  | X |
| **Control** | 61-70 | w | 4 | Pneumonia |  | X |
| **Control** | 41-50 | m | 5 | Transection of spinal cord |  | X |
| **Control** | 71-80 | m | 0 | Suffocation by hanging |  | X |
| **Control** | 71-80 | w | 0 | Traumatic brain injury and suffocation |  | X |

PMI = *post-mortem* interval

**Extended data table 2.** Patients’ demographics for analyses of clinical data.

|  | **Mild** | **Moderate** | **Severe** | **Critical** | **Lethal** |
| --- | --- | --- | --- | --- | --- |
| **N**<br>(% female) | 118 (47) | 48 (69) | 55 (42) | 47 (53) | 55 (64) |
| **Age, mean**<br>**(SD)** | 46.84 (18.8) | 53.69 (17) | 67.67<br>(13.54) | 60.51<br>(15.85) | 70.73 (12.52) |
| **Respiratory rate**<br>**(per minute)** | 13.74 (1.63) | 14.18 (1.67) | 14.76<br>(2.27) | 16.18<br>(2.87) | 14.85 (3.25) |
| **CRP**<br>**(mg L<sup>-1</sup>)** | 16.59 (25.51) | 39.53 (37.67) | 88.05<br>(77.17) | 136.13<br>(105.16) | 128.06<br>(96.01) |
| **Lactate**<br>**(mmol L<sup>-1</sup>)** | 1.28 (0.6) | 1.33 (0.6) | 1.55 (0.7) | 1.51 (0.45) | 2.35 (1.45) |
| **Leukocytes**<br>**(× 10<sup>6</sup> mL<sup>-1</sup>)** | 6.9 (2.63) | 6.35 (2.59) | 7 (3.34) | 10.15<br>(5.77) | 12.47 (9.38) |
| **Peripheral O<sub>2</sub> saturation**<br>**(%)** | 97.28 (2.32) | 95.67 (2.84) | 93.63<br>(3.66) | 93.18<br>(6.65) | 93.31 (6.47) |
| **Venous CO<sub>2</sub> saturation**<br>**(%)** | 45.14 (7.18) | 43.16 (6.49) | 42.9 (5.61) | 43.63<br>(7.01) | 43.96 (8.5) |
| **pH** | 7.4 (0.05) | 7.39 (0.05) | 7.4 (0.06) | 7.37 (0.07) | 7.37 (0.09) |
| **Venous O<sub>2</sub> saturation**<br>**(%)** | 37.77 (22.58) | 38.73 (27.12) | 36.4<br>(17.56) | 47.84<br>(31.62) | 40.65 (37.38) |
| **Heart rate**<br>**(per minute)** | 84.51 (21.13) | 88.09 (12.02) | 87.2<br>(16.21) | 84.16<br>(19.44) | 90.85 (21.63) |
| **Temperature**<br>**(°C)** | 36.64 (0.77) | 37.08 (0.82) | 37.04<br>(0.93) | 36.74<br>(0.88) | 36.92 (1.23) |

**Extended data table 3.** Odds ratio analysis for prediction of lethality in critical COVID-19.

|  | **Odds ratio** | **P-value** | **CI<sub>95%</sub> low** | **CI<sub>95%</sub> high** |
| --- | --- | --- | --- | --- |
| **Peripheral O<sub>2</sub> saturation (%)** | 1.06 | 0.28 | 0.95 | 1.18 |
| **Respiratory rate (per minute)** | 0.9 | 0.03 | 0.82 | 0.99 |
| **CRP (mg L<sup>-1</sup>)** | 1.06 | 0.24 | 0.96 | 1.16 |
| **Leukocytes (× 10<sup>6</sup> mL<sup>-1</sup>)** | 1.06 | 0.15 | 0.98 | 1.14 |
| **Lactate (mmol L<sup>-1</sup>)** | 1.14 | 0.09 | 0.98 | 1.32 |
| **Venous CO<sub>2</sub> saturation (%)** | 0.96 | 0.53 | 0.85 | 1.09 |
| **pH** | 1.05 | 0.4 | 0.94 | 1.18 |
| **Venous O<sub>2</sub> saturation (%)** | 0.88 | 0.07 | 0.78 | 1.01 |
| **Interleukin-6 (ng mL<sup>-1</sup>)** | 0.95 | 0.11 | 0.89 | 1.01 |

**Extended data table 4.**
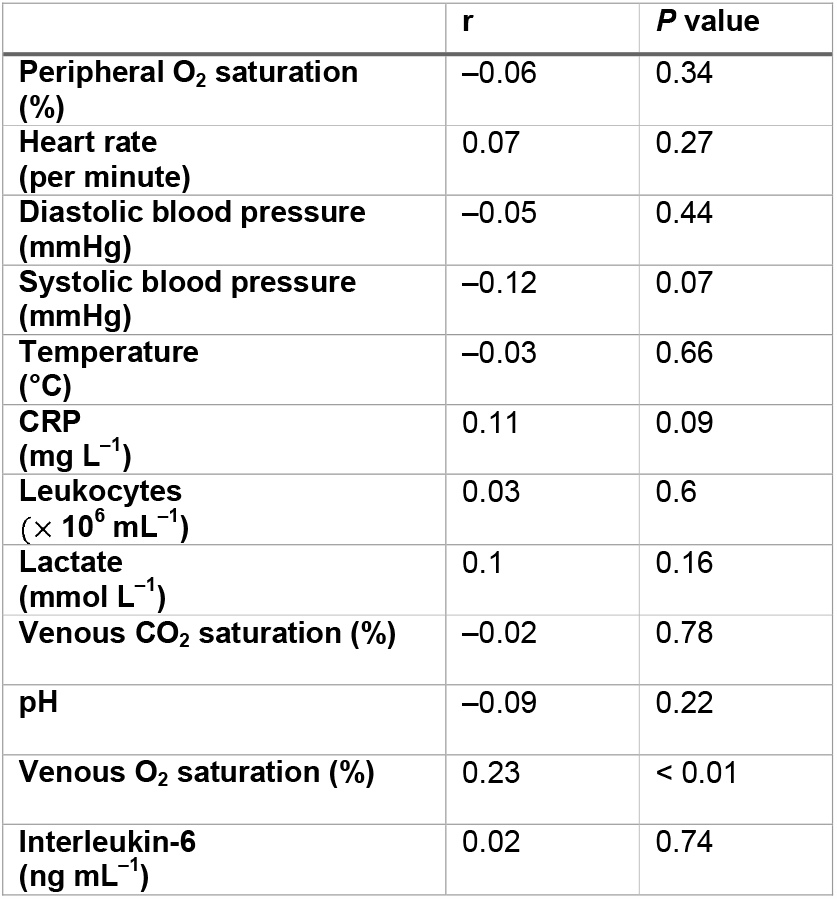
Summary statistics of correlation analyses between respiratory rate and its physiological modulators.

